# A Pilot Study on the use of a Multi-morbidity Index in Patients Receiving Home Parenteral Nutrition. Secondary Analysis of a Quality Improvement Project for Patients on Home Parenteral Nutrition

**DOI:** 10.1101/2023.04.29.23289302

**Authors:** Michael M. Rothkopf, Mohan D. Pant, Zachary S. Rothkopf, Rebecca Brown, Jamie Haselhorst, Francine Gagliardotto, Debbie L. Stevenson, Andrew DePalma, Michael Saracco

**Affiliations:** Rutgers/New Jersey Medical School, Newark, NJ; Eastern Virginia Medical School, Norfolk, VA; St. Luke’s University Hospital, Bethlehem, PA; Amerita, Inc, Greenwood Village, CO; Sea Meadows, LLC, Brick, NJ

## Abstract

**Background:** Home Parenteral Nutrition (HPN) patients often have multiple concomitant medical conditions. A validated multi-morbidity index (MMI) could help determine risks and need of HPN patients. We applied an MMI analysis in a Quality Improvement Project for HPN (QIP-PN) to determine if MMI scoring predicted outcomes.

**Materials and Methods:** Data from 60 HPN patients were analyzed for Cumulative Illness Rating Scale (CIRS) score, PN formula changes, hospitalizations and hospital length of stay (LOS). Density plots were produced to select CIRS score cutoff values. A correlation matrix was drawn showing Spearman correlations among the three variables of interest. The data were then subjected to Kruskal-Wallis and Wilcoxon Rank Sum Tests and Negative Binomial Regression Models to determine if the groups differed significantly.

**Results:** CIRS scores ranged from 9-25 (mean = 17.0 ± 3.85). There were 367 PN formula changes for patients with CIRS ≥17, compared to 297 with CIRS <17. There were 19 hospitalizations for patients with CIRS ≥17 (n=19), compared to 10 with CIRS <17. Total hospital LOS was 122 days for patients with CIRS ≥17 (3.8 days/patient) compared to 100 with CIRS <17 (3.57 days/patient). Results of negative binomial regression indicate that the total hospital LOS was significantly higher in patients with a CIRS ≥17 than CIRS<17 at 5% alpha level.

**Conclusion:** CIRS methodology was well suited for HPN patients. CIRS data on long term HPN patients produced a trend wherein higher multi-morbidity scores were associated with greater hospital LOS. This approach requires further study and validation.

**Key Messages Box:**

1. What is already known on this topic? The risk of HPN complications is known to relate to patient demographics, length of functional bowel, duration of HPN therapy and macronutrient content. A validated method for determining multi-morbidity in HPN patients may be beneficial in determining the needs of and risk for each HPN patient.
2. What does this study add? The study explores the role of measuring multi-morbidity scoring in HPN care to determine the risk of adverse effects during therapy. We found a correlation between multi-morbidity and greater length of stay when HPN patients are readmitted to the hospital. If confirmed, such a correlation could help providers determine how to allocate resources during HPN care.
3. How might this study affect research, practice or policy? The study suggests that using validated multi-morbidity index for HPN patients could help healthcare providers and payors allocate resources based on risk analysis.

## Introduction

HPN patients often have concomitant medical conditions that impact their care and confound outcomes. The existence of multi-morbidity in HPN patients may increase their risk for serious complications (i.e., catheter-related bloodstream infections, venous thrombosis, metabolic imbalances, bone disease, kidney stones, and liver disease). However, research has yet to address the role of multi-morbidity in relation to HPN risk. This pilot study was intended to assess the value of measuring multi-morbidity in patients receiving HPN.

The risk of HPN complications varies widely. Previous studies have suggested a relationship to patient demographics, length of functional bowel, duration of HPN therapy and macronutrient content (1, 2). These risks can be reduced by careful monitoring and patient education (3).

An accurate, validated method for determining multi-morbidity in HPN patients would be beneficial for practitioners, researchers and payors. A multi-morbidity index (MMI) could be utilized to better determine the needs of each HPN patient. It could improve communication between providers, better allocate patient care resources and more clearly communicate the complexity of care to payors. From a research perspective, a validated HPN multi-morbidity index could assist in comparisons of patient outcomes, utilization and quality of life (QOL) analysis.

We previously studied the effect of a physician nutrition expert (PNE) led multidisciplinary nutritional support team (MNST) on a Quality Improvement Project for HPN (QIP-PN) (4). In that original study, we showed that intervention from an MNST produced measurable improvements in the care of long-term HPN patients. MNST input improved patients’ self-assessed overall health, while reducing adverse outcomes, rehospitalization and hospital LOS. In the current study we performed a secondary analysis to explore the options for measuring multi-morbidity of patients in our previous report. In this paper, we describe the results of a pilot study on measuring multi-morbidity in HPN patients.

### Choice of Multi-Morbidity Index (MMI)

There are >30 current methods for the measurement of multi-morbidity with no universally agreed upon MMI tool (29). Most MMIs utilize disease counts (5–18). Others use diagnostic groups/clusters or drug counts (19–26), while one index used physiologic data from pulmonary function testing, imaging and laboratory results (27).

For this study, we eliminated consideration of MMIs based on disease counts, drug counts and physiologic parameters because they were deemed less applicable to HPN population. We also sought to a tool that was not geriatric focused, as most HPN patients are <65 years of age (28). We reviewed studies that reported using the Charlson Comorbidity Index (CCI) as an MMI for parenteral nutrition patients (29–31) and utilized the Stirland guide (32) to assist in selecting a MMI tool. At the conclusion of this process, we selected the Cumulative Illness Rating Scale (CIRS) as the MMI tool for this study.

CIRS is a well-validated and reliable index (33). It has been shown to be useful for studies determining risks of mortality, re-admission, or future morbidity (34). Of particular interest, the CIRS can be used to quantify chronic disease burden and act as a prognosticator for patient outcome (35).

The CIRS index determines risk by rating the severity of comorbidities in 15 body systems: cardiac, vascular, hematological, respiratory, otorhinolaryngological, ophthalmological, upper gastrointestinal, lower gastrointestinal, hepatic and pancreatic, renal, genitourinary, musculoskeletal/tegmental, neurologic, endocrine/metabolic/breast and psychiatric. A weighted severity score in each system ranges from 0 to 4.

A value of 0 denoted no problem, healed or minor injuries, past childhood illnesses, minor surgery, uncomplicated healed fractures and other past problems which healed without sequela. A value of 1 denotes current medical conditions that caused mild discomfort or disability, or with occasional exacerbations. A value of 1 could also be used to indicate conditions that have a minor impact on morbidity and medical problems that are not currently active but were significant problems in the past. A value of 2 denotes current medical conditions that require daily treatment or first line therapy. A value of 2 could also be used to indicate conditions with a moderate disability or morbidity (36).

A value of 3 denotes current medical conditions that are chronic and/or not controlled with first line therapy. Such conditions would be expected to produce significant disability. A value of 4 denotes current medical conditions that are severe. This included conditions that require immediate treatment, involve organ failure, severe sensory impairment, severely reduced quality of life and severe impairment in function (37).

For malignancies: A value of 1 denoted cancer diagnosed in the remote past without evidence of recurrence or sequela in the past 10 years. A value of 1 could also be used to indicate skin cancer operated in the past without major sequela, other than melanoma. A value of 2 denotes no evidence of recurrence or sequela in the past 5 years. A value of 3 denotes the use of chemotherapy, radiation or hormonal therapy in the past 5 years. A value of 4 denotes recurrent malignancy or metastasis (other than to lymph glands) or palliative treatment stage (38).

The weighted severity ratings are observer dependent. There is no standardized method regarding what constitutes a 0 vs 4. Because of observer variability, there are concerns regarding the reproducibility of CIRS scoring. Some studies have found that resultant CIRS scores for a patient can differ based on the user’s background (e.g., pulmonologist vs oncologist) (ƒ).

However, the reason for these discordances can be as simple as one user overlooking a minor comorbidity while the other gives more weight to that same comorbidity. In this pilot study, we controlled for this discrepancy by using a multidisciplinary team to score CIRS on our patients.

## Materials and Methods

### Research Question

This pilot study was intended to answer a research question regarding the value of measuring multi-morbidity in patients receiving HPN. Specifically, does multi-morbidity translate into greater resource utilization or increased adverse outcomes in HPN care? Further, can quantification of multi-morbidity help predict the need for additional resources for specific patients? Lastly, can multi-morbidity indexes be a component of a more comprehensive risk assessment tool for HPN care?

This pilot study was conducted as a secondary analysis of data from the quality improvement project of patients on home PN (QIP-PN) (4). The protocol was granted Institutional Review Board (IRB) exemption under NIH guidelines (45CFR 46.104(d)(2)) by the Western IRB on September 4, 2019.

### Study groups

The QIP-PN study was performed at Amerita, Inc, a national home infusion organization (Amerita). All HPN patients treated by Amerita branches were considered for the study. To achieve the desired patient enrollment, 7 branches were selected as “study branches” based upon their volume of long-term HPN (>90 days of therapy) cases. The original QIP-PN study was divided into 3 phases for data review (phases 1a and 1b), observation (phase 2) and intervention (phase 3) (Figure 1).

**Figure 1.**
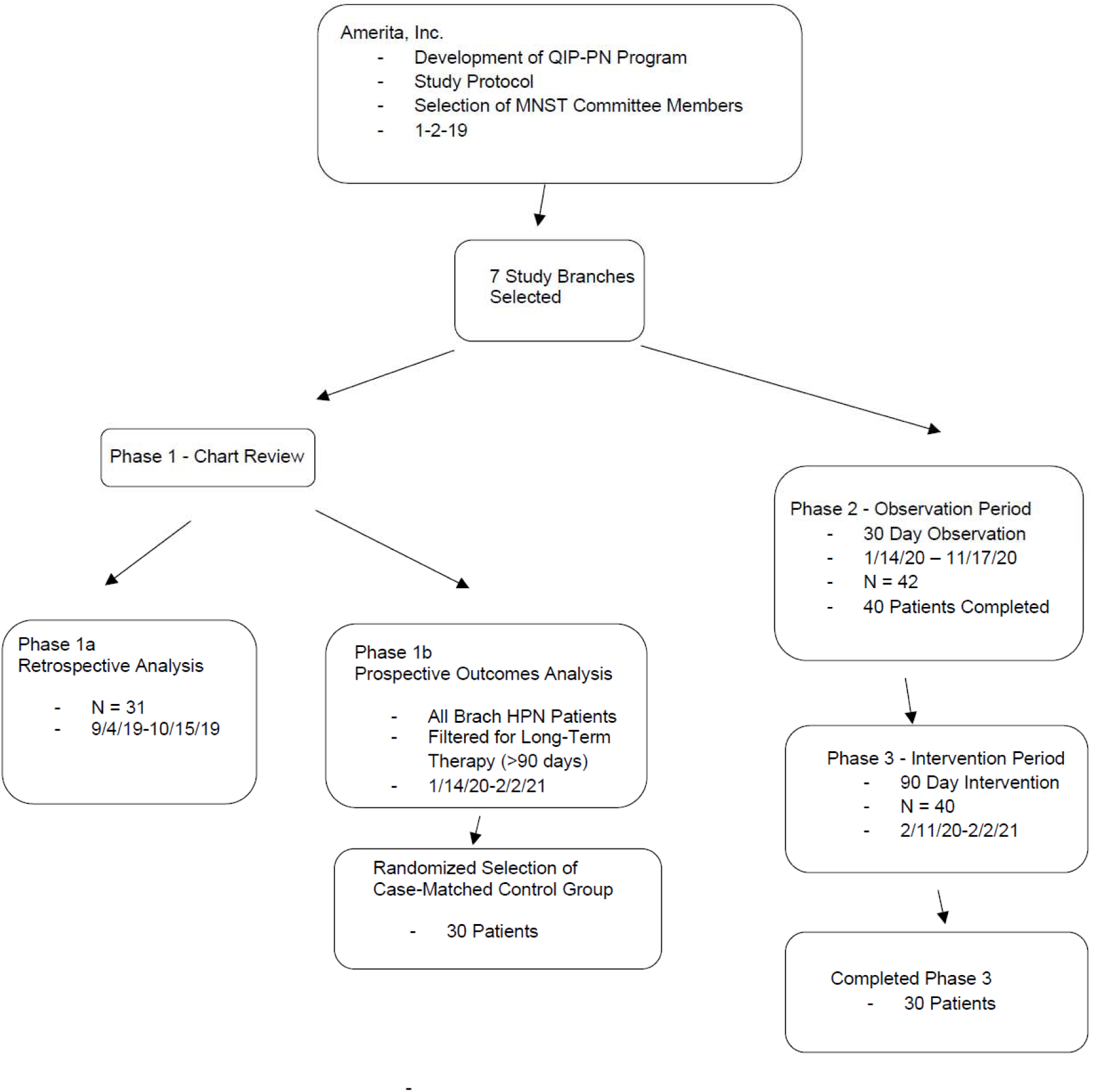
Flowsheet of patient selection and participation in the Amerita QIP-PN Study

Inclusion and exclusion criteria: All long-term adult HPN patients treated at study branches were offered study participation. The inclusion criteria was not limited to diagnosis, age, gender or length of HPN therapy. Patients were excluded from study if they did not sign the study consent form. Patients were also excluded from study if their treating physician did not sign the study participation agreements. 75 HPN patients signed consent forms. Treating physician study participation agreements were signed for 42 patients who comprised the study group, 30 of whom completed phase 3 (Figure 1). A random sample of 30 long-term HPN patients treated at the study branches comprised the case-matched control group (control).

### Study Interventions

MNST meetings discussed each patient case in an “open-book” environment. That is, MNST members (employees or consultants to the HPN Company) had access to the patient’s electronic clinical chart when cases were discussed. Recommendations for PN formula changes, laboratory studies, infusion cycle, catheter care and patient monitoring were made by the MNST and recorded. Recommendations for HPN care modification were provided to the patient’s treating physician by an Amerita branch clinician. HPN care parameters and adverse outcomes were recorded.

### Multi-morbidity Assessment

MNST members received training on the CIRS rating scale, using the method according to DeGroot et al. (35). During each patient’s initial case presentation, MNST members analyzed their comorbidities. At least one MNST member had full access to the electronic medical record during the CIRS scoring sessions. This included hospital discharge notes, homecare admission notes, regular physician and nursing notes, medication reconciliations, emergency department visits, treatment plans, care summaries and other available data. At least one MNST member had direct management experience with the patient. Each patient was presented to the MNST with a brief history of their case and HPN care at their initial case review. A tally of known conditions in each of 15 body systems was performed. Each system received a weighted score of 0-4, as described above. A CIRS-specific spreadsheet was developed which calculated the total CIRS score for each patient and control (Figure 2). Performance of the CIRS scoring analysis took approximately 30 minutes per case.

**Figure 2.**
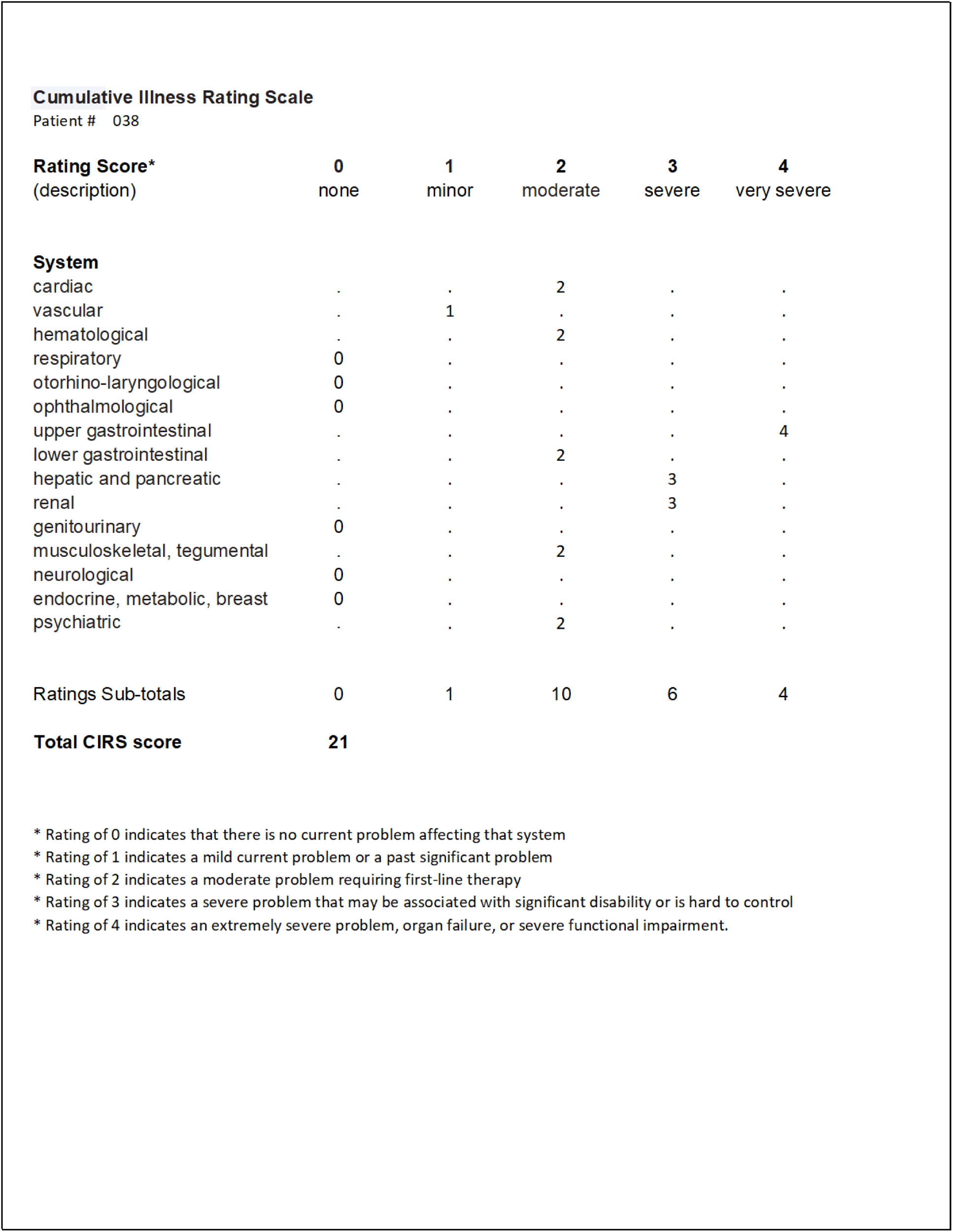
Example of CIRS Rating Scale for Patient # 038.

During the 90-day intervention period, weekly telephonic MNST meetings were held as case conferences via indirect chart review. In the control group no recommendations were made by MNST members. In the study group, MNST recommendations were limited to those that related directly to the HPN process. This included macronutrient adjustments to conform to recommended guidelines, the addition or subtraction of micronutrients based on clinical and lab findings, changes in PN volume and rate based on patient responses, laboratory testing to conform to published standards and the frequency of home nursing and clinic visits.

### Statistical Analysis

To examine the value of MMI in HPN, we elected to commingle the data of the study and case-matched controls. This was feasible because each patient in both groups had undergone CIRS scoring. Another rationale for comingling the data of the two groups was because these two groups were not statistically significantly different (see Table 7) at the 5% significance level. We used a 5% level of significance (alpha level) for all inferential statistical tests. In the comingled group of 60 patients, we had data on the number of total parenteral nutrition (TPN) formula changes, hospitalization rates and length of stay for all patients. We used CIRS score of 17 as a cutoff score for creating two groups CIRS <17 and CIRS ≥17 because it is the mean score of 60 participants in the comingled data and we considered this mean score to serve as a balancing point (like a fulcrum) in the distribution of CIRS score. As indicated by the results in Table 7, the study and control groups were statistically similar. Therefore, these groups were combined.

We used a nonparametric statistical approach for comparison between the two groups (CIRS <17 and CIRS ≥17) on the variables of interest, which are Formula Changes, Hospitalization, and Total LOS. The two groups (CIRS <17 and CIRS ≥17) were formed based on the criterion that mean CIRS score of 17 (of the combined data of 60 participants) would serve as the balancing point (like a fulcrum). Density plots were produced to show approximate similarity in the distributions of the variables of interest between the two CIRS-based groups. A correlation matrix was drawn showing Spearman correlations among the three variables of interest (Figure 6) to assess the degree of association among these variables. The data were then subjected to Wilcoxon rank sum tests and negative binomial regression model (tables 7-9). Statistical analysis included computation of descriptive statistics in the form of Mean (M), Standard Deviation (SD), Minimum (Min), Median, Interquartile Range (IQR), Maximum (Max) to summarize the three variables of interest with count data. Additionally, the data were subjected to Wilcoxon Rank Sum Tests and Negative Binomial Regression Models to determine if the groups differed significantly on the variables of interest. Negative binomial regression model is considered appropriate when the data are over-dispersed, that is, when conditional variance of response variable is greater than the conditional mean (39). In this study, the variables of formula changes, hospitalizations, and total LOS are all over-dispersed. Therefore, negative binomial regression model is a suitable approach.

### Outcomes

The outcomes studied consisted of the number of PN formula changes, hospitalizations and hospital length of stay for each effected individual with the comingled group of study and control patients.

## Results

The average age of 30 study patients was 57.43 years. Twenty-two were female (73.3%) and 8 were male (26.7%). In the 30 case-matched controls, the average age was 54.9 years. Twenty-four (80.0%) were female and 6 (20.0%) were male. The CIRS score ranged from 10-25 with a mean of 17.4 ± 3 among study patients and from 9 to 24 among case-matched controls with a mean of 16.5 ± 3.82 (Table 1). The patients’ diagnostic information is presented in Table 2.

**Table 1.**
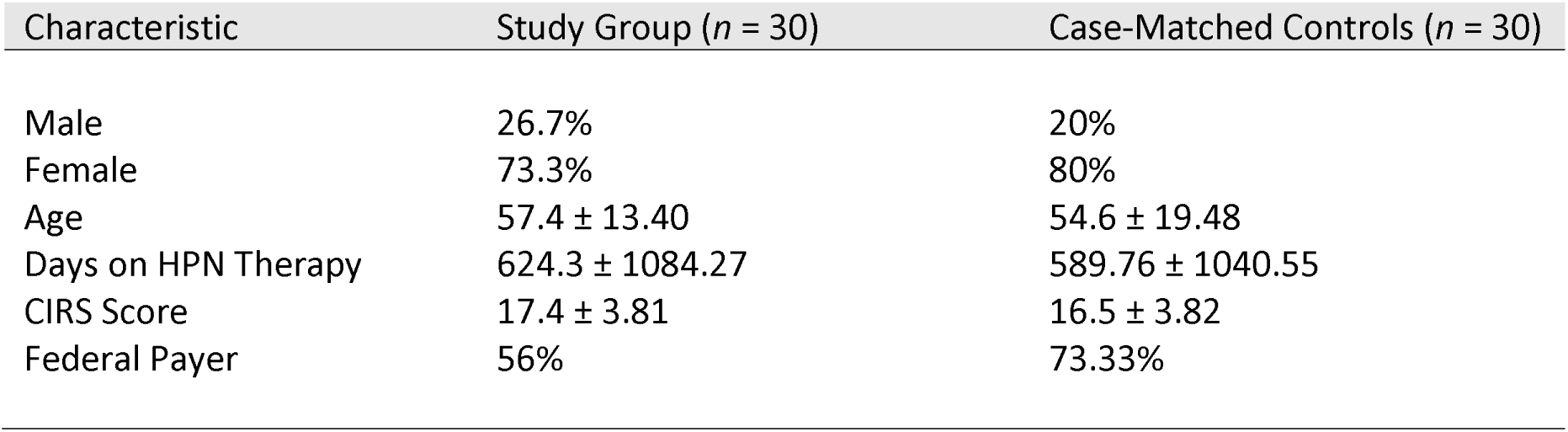
Demographic characteristics of study patients and case-matched controls.

**Table 2.**
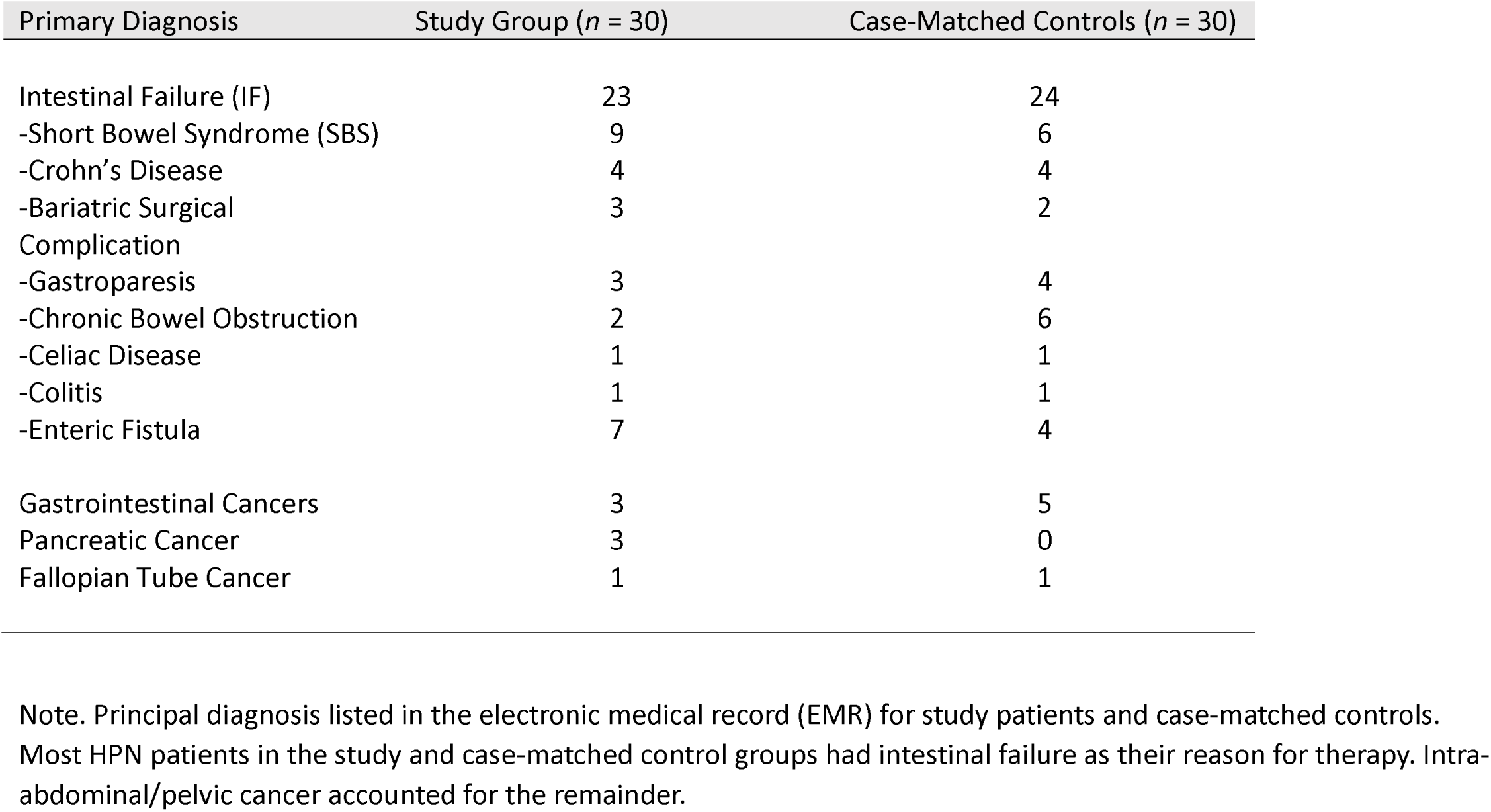
Diagnostic categories of study patients and controls.

| Primary Diagnosis | Study Group (*n* = 30) | Case-Matched Controls (*n* = 30) |
| --- | --- | --- |
| Intestinal Failure (IF) | 23 | 24 |
| -Short Bowel Syndrome (SBS) | 9 | 6 |
| -Crohn's Disease | 4 | 4 |
| -Bariatric Surgical | 3 | 2 |
| Complication |  |  |
| -Gastroparesis | 3 | 4 |
| -Chronic Bowel Obstruction | 2 | 6 |
| -Celiac Disease | 1 | 1 |
| -Colitis | 1 | 1 |
| -Enteric Fistula | 7 | 4 |
| Gastrointestinal Cancers | 3 | 5 |
| Pancreatic Cancer | 3 | 0 |
| Fallopian Tube Cancer | 1 | 1 |

Note. Principal diagnosis listed in the electronic medical record (EMR) for study patients and case-matched controls. Most HPN patients in the study and case-matched control groups had intestinal failure as their reason for therapy. Intra-abdominal/pelvic cancer accounted for the remainder.

Ninety (90)-day readmission and hospitalization rate per study group patient were 0.13 and 0.37 respectively. The rates in the control group were 0.23 and 0.67. The average length of hospitalization was 6.27 (days) in the study group and 7.65 in controls (Table 3).

**Table 3.** Adverse outcomes in study patients and controls.

| Outcomes | Study Group (*n* = 30) | Case-Matched Controls (*n* = 30) |
| --- | --- | --- |
| Total Adverse Outcomes | 10 | 16 |
| Access Device Occlusion | 1 | 0 |
| Emergency Room Visits | 2 | 2 |
| Unplanned Hospitalization | 7 | 14 |
| PN-related Hospitalization | 3 | 3 |
| - PN-unrelated Hospitalization | 4 | 11 |
| - Total Hospital Admissions | 11 | 20 |
| Single Hospital Admission | 3 | 8 |
| - Double Hospital Admission | 4 | 5 |
| - Triple Hospital Admission | 0 | 2 |
| - Readmission Rate | 0.13 | 0.23 |
| Hospitalization Rate per Patient | 0.37 | 0.67 |
| Total Hospital Length of Stay (LOS) Days | 69 | 153 |
| Average LOS Day | 6.27 | 7.65 |

Note. Study patients had fewer total adverse outcomes and unplanned hospitalization than case-matched controls. Emergency department use was similar in both groups. Study group patients had a lower hospitalization rate, readmission rate, total LOS and average LOS than case-matched controls. Note: LOS = length of stay, PN = parenteral nutrition.

After the study group and case-matched controls datasets were comingled, the mean CIRS score was 17.0 ± 3.85. This permitted us to separate patients into those with CIRS scores above or below the mean. Roughly half of the patients fell into each group: 32 (53%) above and 28 (47%) below the mean. Descriptive statistics for variables between the two groups (CIRS <17 and CIRS ≥17) did not show significant differences between the groups (Table 4). Also, the Wilcoxon Rank Sum tests conducted separately for each variable (Formula Changes, Hospitalizations, and Total LOS) to determine the difference between two groups did not yield significant results (see Tables 7 and 8 for the results of Wilcoxon Rank Sum tests). The negative binomial regression model (appropriate for count response variable) for modeling Hospitalizations as a function of CIRS Group (CIRS <17 vs CIRS ≥17) and Formula Changes yielded significant results. Specifically, presented in Table 6 are the results of this model. From Table 6, the CIRS ≥17 group interacted with Formula Changes was significantly different from the CIRS <17 group interacted with Formula Changes. This implies that Hospitalizations were significantly different between two groups (CIRS <17 and CIRS ≥17) across all levels of formula changes.

**Table 4:** Descriptive Statistics for Variables between the two Groups (CIRS ≥ 17 and CIRS < 17)

| Variable | M | SD | Min | Median | IQR | Max | N |
| --- | --- | --- | --- | --- | --- | --- | --- |
| Formula Changes |  |  |  |  |  |  |  |
| CIRS $\geq 17$ | 11.50 | 11.00 | 0 | 9.50 | 12.00 | 41 | 32 |
| CIRS $< 17$ | 10.60 | 9.62 | 0 | 8.00 | 11.80 | 39 | 28 |
| Hospitalizations |  |  |  |  |  |  |  |
| CIRS $\geq 17$ | 0.59 | 0.98 | 0 | 0 | 1.00 | 3 | 32 |
| CIRS $< 17$ | 0.43 | 0.63 | 0 | 0 | 1.00 | 2 | 28 |
| Total LOS |  |  |  |  |  |  |  |
| CIRS $\geq 17$ | 3.81 | 7.28 | 0 | 0 | 6 | 27 | 32 |
| CIRS $< 17$ | 3.57 | 7.70 | 0 | 0 | 2.25 | 34 | 28 |

Note. No significant demographic differences were identified between the groups. CIRS = cumulative illness rating scale

**Table 5.** Homecare utilization, hospitalization and hospital length of stay during a 90-day study period.

| | Comingled<br>CIRS $< 17$ | Study<br>CIRS $< 17$ | Control<br>CIRS $< 17$ | Comingled<br>CIRS $\geq 17$ | Study<br>CIRS $\geq 17$ | Control<br>CIRS $\geq 17$ |
| --- | --- | --- | --- | --- | --- | --- |
| Patients | 28 | 12 | 16 | 32 | 18 | 14 |
| Number of PN<br>Formula Changes | 297 | 143 | 154 | 367 | 230 | 137 |
| Total<br>Hospitalizations | 12 | 2 | 10 | 19 | 9 | 10 |
| Hospital Length of<br>Stay | 100 | 21 | 79 | 122 | 48 | 74 |

Note. Comingled data of study patients and case-matched controls, stratified by CIRS scores.

**Table 6.** Negative Binomial Regression Model for Regressing Hospitalizations on CIRS Group and Formula Changes.

|  | Estimate | Std. Error | z value | Pr(>\|z\|) |
| --- | --- | --- | --- | --- |
| CIRS ≥ 17:Formula_Changes | 0.05508 | 0.01581 | 3.484 | 0.000495 *** |
| CIRS < 17:Formula_Changes | 0.03002 | 0.02365 | 1.270 | 0.204238 |

Note: Based on the Results shown in the Coefficients table, the number of hospitalizations is significantly different, at the 5% alpha level, for the group CIRS ≥ 17 interacted with Formula Changes compared with the group CIRS < 17 interacted with Formula Changes.

**Table 7:**
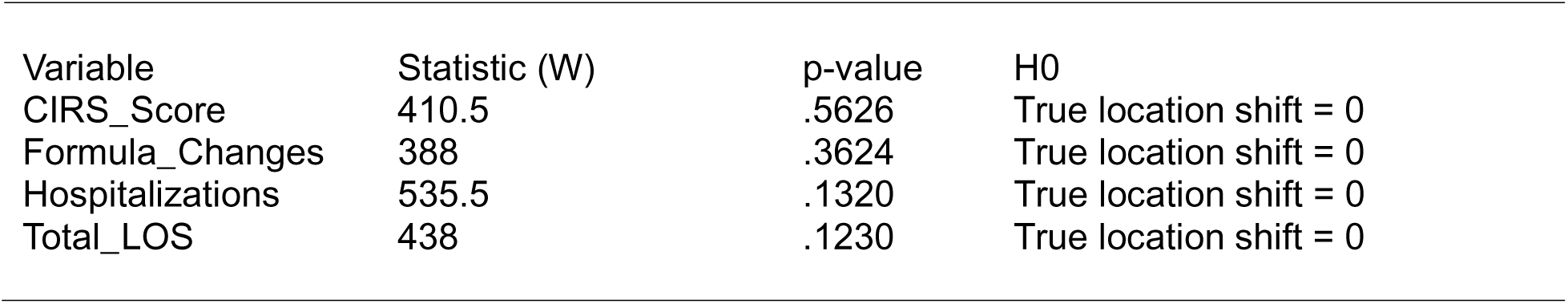
Wilcoxon Rank Sum Test between the two Study and Control Groups.

**Table 8:** Wilcoxon Rank Sum Test between the two Groups CIRS ≥ 17 and CIRS < 17.

| Variable | Statistic (W) | p-value | H0 |
| --- | --- | --- | --- |
| Formula_Changes | 449.5 | .9882 | True location shift = 0 |
| Hospitalizations | 455 | .9081 | True location shift = 0 |
| Total_LOS | 439 | .8806 | True location shift = 0 |

Data were compared for the number of PN formula changes, total hospitalizations, and hospital length of stay during the 90-day study period between patients with a CIRS score above or below the mean (Figures 3-5). During the study period, PN formula changes differed for patients with CIRS equal to or above the mean (n=367) compared to those with CIRS scores below the mean (n=297). Total hospitalizations differed for patients with CIRS scores equal to or above the mean (n=19) compared to those with CIRS scores below the mean (n=10). Total hospital LOS differed for patients with CIRS scores equal to or above the mean (n=122 days; 3.8/patient) compared to those with CIRS scores below the mean (n=100 days; 3.57/patient) (Table 5). A correlation matrix was drawn showing Spearman correlations among the three variables of interest (Figure 6). Wilcoxon Rank Sum Tests and Negative Binomial Regression Models revealed that the number of hospitalizations was significantly different for the group CIRS ≥17 interacted with Formula Changes compared with the group CIRS <17 interacted with Formula Changes (Table 6). This implies that Hospitalizations were significantly different between two groups (CIRS <17 and CIRS ≥17) across all levels of formula changes. Inspection of Table 7 indicates that the study and control groups were not statistically significantly different on each of the variables of CIRS score, Formula changes, Hospitalization, and Total length of stay (LOS). Likewise, inspection of Table 8 indicates that the CIRS-based groups (CIRS <17 and CIRS ≥17) were not statistically significantly different on each of the variables of Formula changes, Hospitalization, and Total LOS. The variable CIRS score was normally distributed (Shapiro-Wilk test: W = 0.98, p = .3796). Therefore, we ran a two-sample t-test with equality of variance assumption. The results as reported in Table 9 were statistically significant, *t*(*df* = 58) = 10.91, p < .0001, implying that the mean CIRS scores of the two groups CIRS <17 and CIRS ≥17were significantly different in the population, which is an obvious fact.

**Figure 3:**
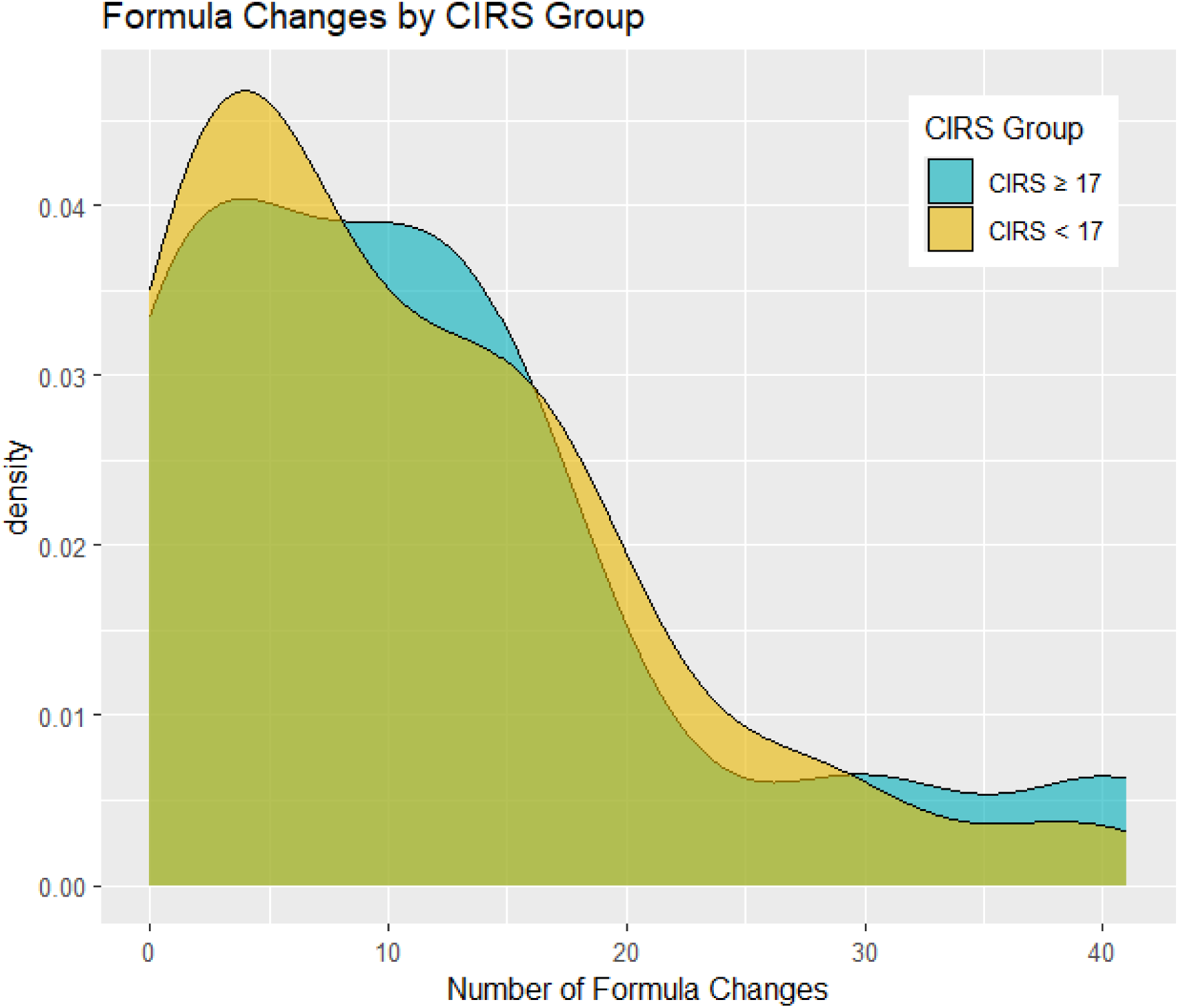
Density Plot of Formula Changes for the two Groups (CIRS *≥* 17 and CIRS < 17)

**Figure 4:**
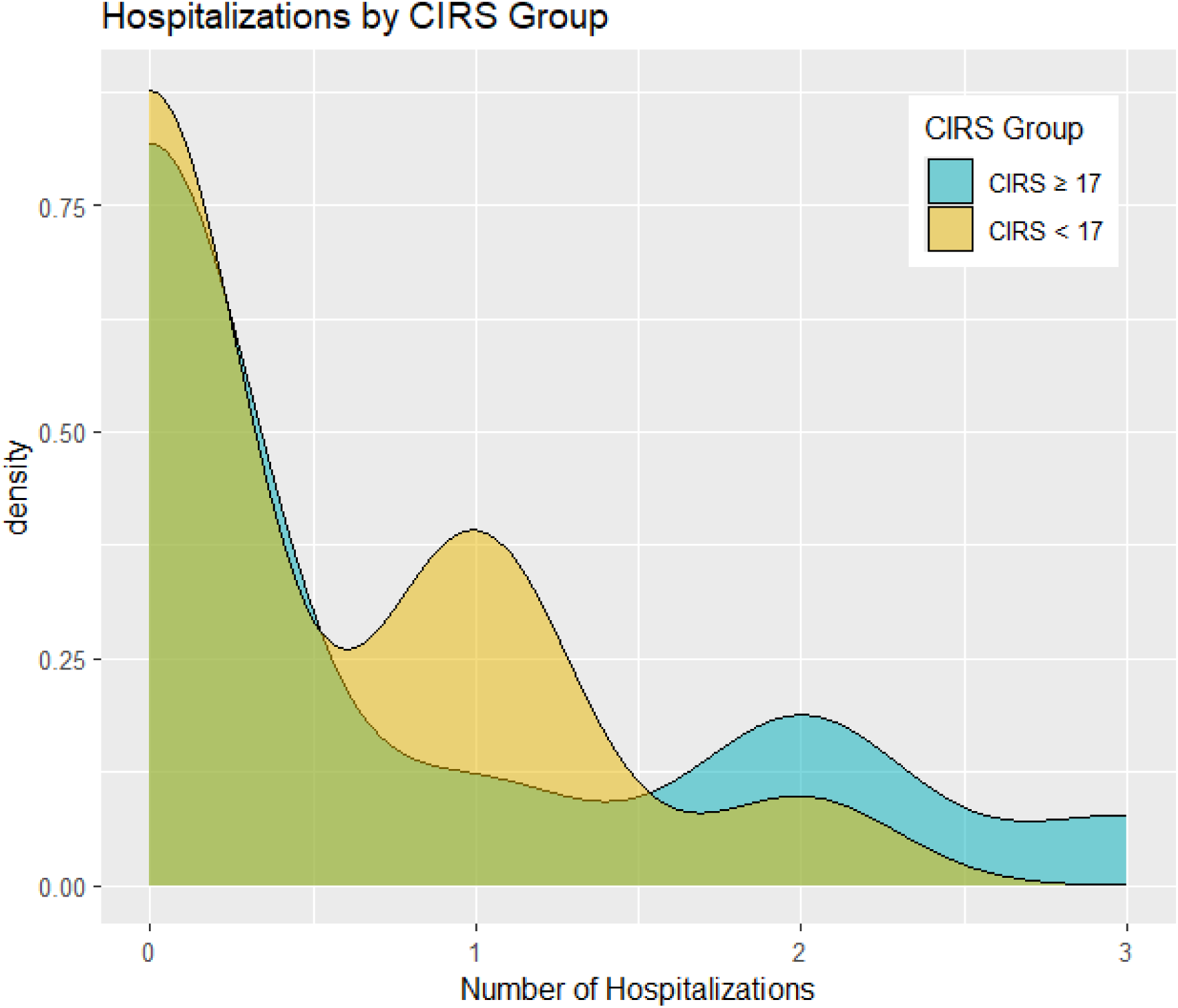
Density Plot of Hospitalizations for the two Groups (CIRS *≥* 17 and CIRS < 17)

**Figure 5:**
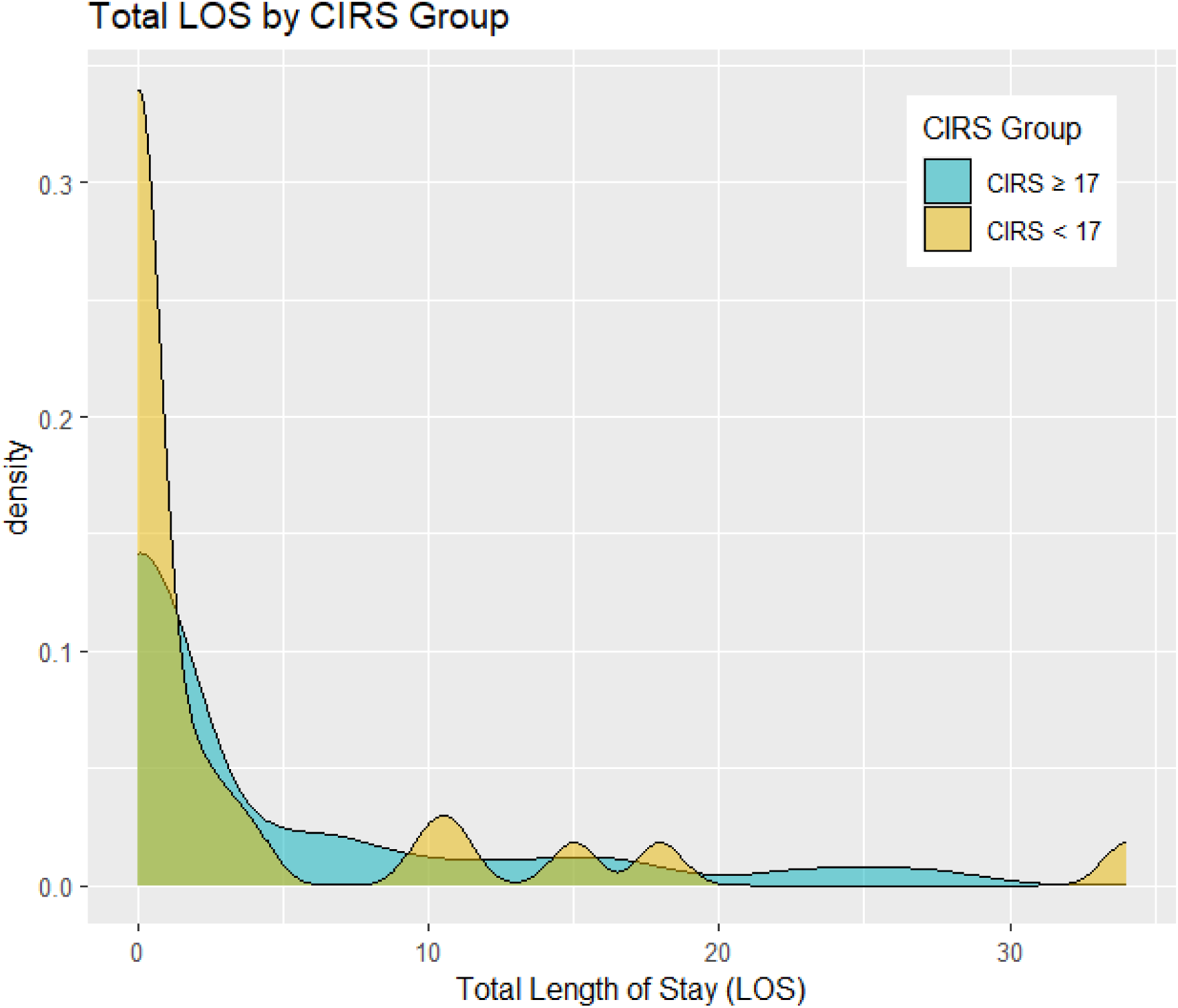
Density Plot of Total Length of Stay (LOS) for the two Groups (CIRS *≥* 17 and CIRS < 17)

**Figure 6:**
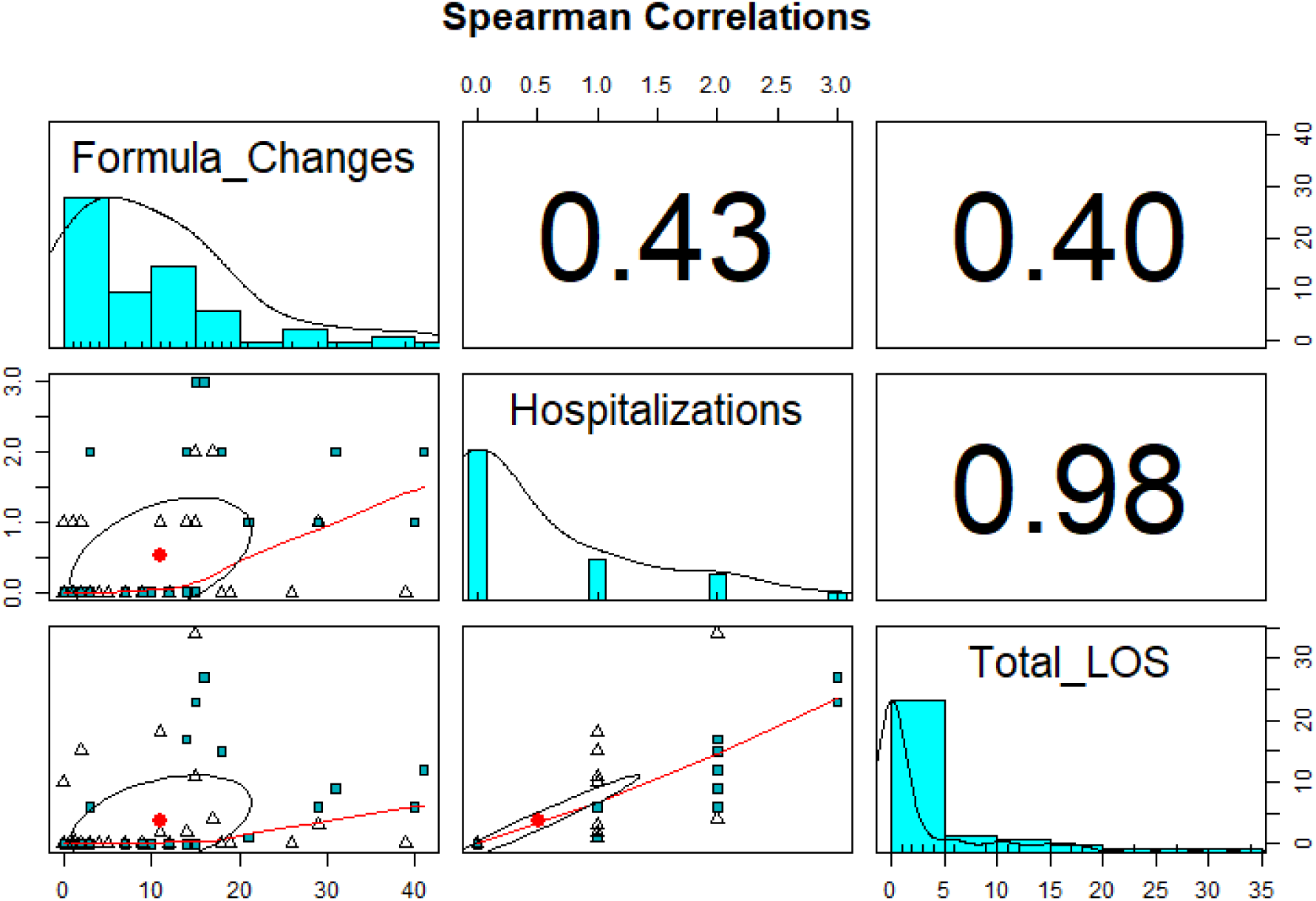
Correlation Matrix Plot Showing Spearman Correlations among the three Variables of Interest (Formula Changes, Hospitalizations, and Total LOS) for the entire Sample of n = 60.

**Table 9:**
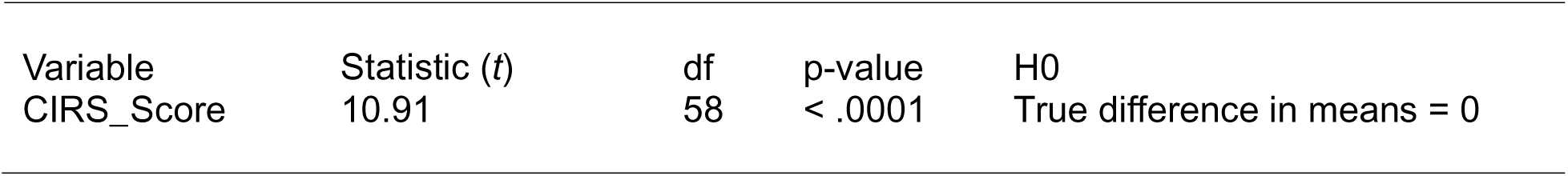
Two-sample t-test between the two Groups CIRS ≥ 17 and CIRS < 17.

| Variable | Statistic (t) | df | p-value | H0 |
| --- | --- | --- | --- | --- |
| CIRS_Score | 10.91 | 58 | < .0001 | True difference in means = 0 |

Inspection of Table 7 indicates that the study and control groups were not statistically significantly different on each of the variables of CIRS score, Formula changes, Hospitalization, and Total length of stay (LOS). Likewise, inspection of Table 8 indicates that the CIRS-based groups (CIRS <17 and CIRS ≥17) were not statistically significantly different on each of the variables of Formula changes, Hospitalization, and Total LOS. The variable CIRS score was normally distributed (Shapiro-Wilk test: W = 0.98, p = .3796). Therefore, we ran a two-sample t-test with equality of variance assumption. The results as reported in Table 9 were statistically significant, *t*(*df* = 58) = 10.91, p < .0001, implying that the mean CIRS scores of the two groups CIRS <17 and CIRS ≥17 were significantly different in the population, which is an obvious fact.

## Discussion

In this paper, we explored the possibility of utilizing multi-morbidity tools to help assess and compare patients receiving HPN therapy. We sought to determine whether measurement of multi-morbidity could identify patients who would require greater HPN resource utilization.

We utilized the number of parenteral nutrition formula changes as a surrogate marker for HPN resource utilization. Such an approach has validity because the number of PN formula changes reflects patient stability, laboratory results, changes in clinical condition and potential for adverse outcomes.

Furthermore, we sought to determine whether multi-morbidity scores could predict patients who are at higher or lower risk for adverse outcomes in HPN care. For this determination, we examined total hospitalization and length of hospital stay as end points. This approach has validity because patients with more adverse outcomes can be expected to have a higher number of hospitalizations and greater lengths of stay. A literature review of the available methods for multi-morbidity scoring resulted in our determining the applicability of these methods for the HPN population. We then utilized Stirland et al. (29) guide to select the CIRS index as most applicable for HPN care.

The application of a CIRS scoring system to HPN patients yielded some valuable insights. Training of MNST team members on the CIRS methodology was relatively straightforward. CIRS index scoring consumed approximately 30 minutes of MNST time per patient.

The results of this pilot study uncovered a possible relationship between higher multi-morbidity scores and utilization, hospitalization and hospital length of stay. A significant relationship was found in the occurrence of total hospitalizations and elevated CIRS score (Table 6).

Multi-morbidity adds a degree of complexity that can impact the care of patients receiving HPN. It can require more frequent visits, lab work and hospitalizations. The degree of multi-morbidity may place additional responsibilities on the care team as they factor the interactions of non-nutritional conditions and treatments with the PN therapy. HPN outcomes research suffers from a lack of standardization for multi-morbidity measurement HPN patients which often leads to a comparison of “apples and oranges” (39).

Our pilot study applied the CIRS method to a group of HPN patients who were already under study as part of a quality improvement initiative (QIP-PN). We found the CIRS method was well suited for HPN patient assessment. It was easy to learn and perform. It provided a single variable which could be used as a surrogate of the patient’s multi-morbidity.

When the CIRS data on our long term HPN patients was analyzed, a trend emerged wherein a higher CIRS score was associated with greater resource HPN utilization and adverse events. This approach requires further study and validation.

It is important to acknowledge that there were several limitations to the current study. The size of our study group was small, only 60 patients in total. In addition, our data also contained some missing values. A larger study population may be needed to fully determine the value of MMI scoring. Furthermore, the selection of the CIRS method for MMI scoring may not be optimal for the HPN population. Other MMI tools such as the Charlson score may give different results and validity. The Charlson score has been previously utilized for MMI in HPN (40). The surrogate markers we utilized (number of PN formula changes, number of hospitalizations and hospital length of stay) may not be the optimal choice as indicators of outcomes.

On final note, we do not believe that MMI scoring is reimbursable by third-party payers. Therefore, its use may incur some added costs to the HPN organization.

## Data Availability

All data produced in the present study are available upon reasonable request to the authors

## Disclosures

### Financial

None to declare

## Acknowledgments

The authors acknowledge the guidance of Amy Rothkopf, MS, MSN, APN on multi-morbidity index tools in chronic illness.

## Data Sharing

Data described in the manuscript will be made available upon request to the first author, pending application and approval.

